# Amplitude modulation perception and cortical evoked potentials in children with listening difficulties and their typically-developing peers

**DOI:** 10.1101/2023.10.26.23297523

**Authors:** Lauren Petley, Chelsea Blankenship, Lisa L. Hunter, Hannah J. Stewart, Li Lin, David R. Moore

**Affiliations:** Communication Sciences Research Center; Patient Services Research, Cincinnati Children’s Hospital Medical Center, Cincinnati, Ohio. USA; Department of Psychology, Clarkson University, Potsdam, NY. USA; College of Medicine, Otolaryngology and University of Cincinnati, Cincinnati, Ohio. USA; College of Allied Health Sciences, Communication Sciences and Disorders, University of Cincinnati, Cincinnati, Ohio. USA; Department of Psychology, Lancaster University, U.K; Manchester Centre for Audiology and Deafness, University of Manchester, U.K

## Abstract

**Purpose:** Amplitude modulations (AM) are important for speech intelligibility, and deficits in speech intelligibility are a leading source of impairment in childhood listening difficulties (LiD). The present study aimed to explore the relationships between AM perception and speech-in-noise (SiN) comprehension in children and to determine whether deficits in AM processing contribute to childhood LiD. Evoked responses were used to parse the neural origin of AM processing.

**Method:** Forty-one children with LiD and forty-four typically-developing children, ages 8-16 y.o., participated in the study. Behavioral AM depth thresholds were measured at 4 and 40 Hz. SiN tasks included the LiSN-S and a Coordinate Response Measure (CRM)-based task. Evoked responses were obtained during an AM Change detection task using alternations between 4 and 40 Hz, including the N1 of the acoustic change complex, auditory steady-state response (ASSR), P300, and a late positive response (LP). Maturational effects were explored via age correlations.

**Results:** Age correlated with 4 Hz AM thresholds, CRM Separated Talker scores, and N1 amplitude. Age-normed LiSN-S scores obtained without spatial or talker cues correlated with age-corrected 4 Hz AM thresholds and area under the LP curve. CRM Separated Talker scores correlated with AM thresholds and area under the LP curve. Most behavioral measures of AM perception correlated with the SNR and phase coherence of the 40 Hz ASSR. AM Change RT also correlated with area under the LP curve. Children with LiD exhibited deficits with respect to 4 Hz thresholds, AM Change accuracy, and area under the LP curve.

**Conclusions:** The observed relationships between AM perception and SiN performance extend the evidence that modulation perception is important for understanding SiN in childhood. In line with this finding, children with LiD demonstrated poorer performance on some measures of AM perception, but their evoked responses implicated a primarily cognitive deficit.

## Introduction

Listening difficulties (LiD) have recently assumed a central role in hearing science as an umbrella term for various problems, primarily occurring despite clinically normal audiometry (Dillon & Cameron, 2021; Moore, 2018). While hearing is a passive process, listening requires selective attention and the interpretation of auditory input, which can be an effortful process even for speech that is clearly audible (Pichora-Fuller et al., 2016). Thus, listening additionally involves cognitive brain regions, such as the dorsal frontoparietal attention network (Corbetta & Shulman, 2002) and the temporo-frontal language network (Friederici, 2011). These top-down mechanisms can impact early stages of perceptual organization including auditory scene segregation (Elhilali et al., 2009) and perceptual grouping (Davis & Johnsrude, 2007). In sum, listening is considerably more demanding than hearing. The present study explores the interactions between sensation, perception, and cognition that might contribute to LiD.

About half of the adult patients who seek audiological assessment, estimated at 40 million in the USA alone (Edwards, 2020), present with clinically normal audiograms and LiD of enigmatic origin. LiD commonly involves reduced speech intelligibility under challenging listening conditions, such as those involving noisy, rapid, or degraded speech. It encompasses the spectrum of speech perception deficits that can be experienced by both children and adults (Dillon & Cameron, 2021). LiD is related to the clinical construct of auditory processing disorder (APD). However, inconsistencies in the diagnosis of APD have provoked debate regarding whether APD should be used as a diagnostic label (Moore, 2018; Wilson & Arnott, 2013). Many position statements endorse the view that APD arises from disturbed central auditory processing (i.e., abnormal processing at some level of the central auditory nervous system, Moore et al., 2013). There is reason to question the validity of that assertion. For example, several recent studies with pediatric and adolescent samples have highlighted cognitive deficits as major contributing factors in LiD (e.g., McGrath et al., 2023; Pascoinelli et al., 2021; Petley et al., 2021).

Physiological evoked responses have long been endorsed for use in the clinical assessment of APD (Jerger & Musiek, 2000). They can provide valuable insight into the stages of processing that lead from auditory sensation to perception, including the influence of cognition (Joos et al., 2014). For example, responses like the auditory N1 and the auditory steady state response (ASSR) are largely sensory, with source generators in the central auditory nervous system (CANS), though modulation by selective attention is possible (Hillyard et al., 1973; D.-W. Kim et al., 2011; Skosnik et al., 2007; Talsma & Woldorff, 2005). By contrast, positive event-related potentials (ERPs) arising approximately 300 ms from stimulus onset are linked to attention. The most extensively studied among them is the P300, which is evoked by target stimuli and is generally not observed under conditions of inattention (Duncan et al., 2009; Polich, 2007). P300 is a cognitive response; it is neither restricted to auditory stimuli nor generated in the CANS. Its neural sources are poorly understood but, consistent with its link to selective attention, it has been attributed to a network of frontal and temporal/parietal regions (Polich, 2007).

In the context of LiD, evoked responses could be applied to study any perceptual skill that supports speech comprehension under adverse listening conditions. One crucial skill for speech perception is the ability to accurately perceive amplitude modulations (AM). The importance of AM for speech perception is well-established (Rosen, 1992; Shannon et al., 1995). Analyses of the temporal modulation rate of speech across different languages (such as American English, Chinese, and Swedish) have revealed remarkable similarities in their modulation rates, which generally lie between 2 and 10 Hz (Ding et al., 2017), but can extend up to about 50 Hz (Rosen, 1992).

The temporal modulations of speech are also powerfully related to its intelligibility under challenging listening conditions. For example, envelope periodicity is a major contributor to speech intelligibility in the presence of a competing talker (Christiansen et al., 2013). Indeed, with modulated maskers like natural speech, the envelope of the masker is itself an important factor, since periods of low masker energy provide opportunities for “glimpsing” the target (Festen & Plomp, 1990; Gnansia et al., 2008). Furthermore, electrophysiological measurements suggest that phase-locking of neural oscillations to the amplitude envelope of speech is a crucial mechanism for biasing cortical processing towards the attended stream (Horton et al., 2013). Thus, there is ample evidence that accurate neural representations of AM provide numerous benefits for speech perception, both in quiet and in the presence of noise.

Current theories of how AM is encoded and represented in the auditory system involve both peripheral and central mechanisms. Models of the earliest stages of processing, in the cochlea and cochlear nucleus, decompose acoustic stimuli into half-wave rectified, compressed, and low-pass filtered narrowband signals (Viemeister, 1979; Yang et al., 1992). AM is well-represented in the CANS, all the way to primary auditory cortex (Joris et al., 2004). However, behavioral thresholds for AM detection improve over childhood (Cabrera et al., 2022; Hall & Grose, 1994; Talarico et al., 2007). Consistent with the idea that deficits in modulation perception might be involved in APD, a recent study by Lotfi and colleagues (2020) demonstrated elevated thresholds for detecting spectrotemporal modulation in children with APD across several temporal modulation rates and spectral modulation densities (Lotfi et al., 2020).

Neural synchronization to AM sounds can be measured via the ASSR, which is readily evoked in children and infants and has been used to study the neural mechanisms of conditions related to LiD, such as dyslexia (De Vos et al., 2020). The ASSR is influenced by the audibility of stimuli with sufficient reliability that it is effective for measuring audiometric thresholds (Luts et al., 2004). Thus, it is an effective index of sensory processing for AM stimuli. Similarly to thresholds for AM perception, the ASSR changes over childhood and adolescence, particularly with respect to its amplitude at 40 Hz, which reaches its maximum in early adulthood (Aoyagi et al., 1993; Cho et al., 2015; Rojas et al., 2006). Another evoked response that is valuable for the study of perception is the acoustic change complex (ACC). The ACC is a transient response to a change in a continuous stimulus, such as a change in tonal frequency or intensity, which is composed of an N1 and subsequent P2 component (Martin & Boothroyd, 2000). Unlike the N1 and P2 that are evoked by stimulus onsets, the ACC is correlated with psychometric discrimination thresholds, and has been proposed as an objective index for their measurement (He et al., 2012; J.-R. Kim, 2015).

The N1 component of the ACC that is evoked by changes in AM may reflect the temporal resolution of AM perception (Han & Dimitrijevic, 2015). Elevated AM detection thresholds and smaller, later N1 components elicited by AM change have also been observed in adults with cochlear implants relative to typically-hearing controls, suggesting that the ACC to changes in AM rate may be a useful index of speech perception abilities (Han & Dimitrijevic, 2020). While this research suggests that the ACC to changes in AM rate may be valuable for the study of populations with speech perception deficits, there is a paucity of research demonstrating links between measures of AM and speech perception in children (for examples, see Cabrera et al., 2019; Lotfi et al., 2020), particularly for continuous speech. It is also unknown whether other evoked responses that can be measured using this stimulation protocol, notably the ASSR, vary systematically with AM thresholds in pediatric populations.

The goals of this study were to investigate whether (1) there are relationships between measures of AM and SiN perception in children, (2) evoked responses to AM stimuli correlate with behavioral measures of AM perception in children, and (3) deficits in AM perception are present in children with LiD, as reflected by impaired performance on AM tasks and differences in evoked responses to AM stimuli versus their TD peers. Since late childhood and adolescence are periods of considerable development with respect to auditory perceptual skills (Lopez-Poveda, 2014; Moore et al., 2008), goals (1) and (2) were pursued following examinations of the influence of age, and goal (3) was addressed using age-matched groups to focus on the underlying mechanisms of LiD.

## Method

### Participants

Forty-one children with LiD (8.1 – 15.5 years of age) and forty-four typically developing (TD) children (8.6 – 16.8 years of age) participated in this study. Eligibility, recruitment strategies, and testing procedures were the same as for other reports derived from this research program (Hunter et al., 2020, 2023; D. R. Moore et al., 2020; Petley et al., 2021; Stewart et al., 2022). In brief, the requirements included English as a native language, and the absence of any neurological, psychiatric, or intellectual condition that would hinder test completion. Participants in the TD group additionally could not have a diagnosed developmental delay, or an attention or learning disorder. Information regarding these inclusion criteria and other characteristics such as health background, and sociodemographics were provided by caregivers via a structured background questionnaire. This study was approved by the Institutional Review Board of Cincinnati Children’s Hospital Research Foundation and participants received monetary compensation for their time. Demographics regarding age, sex, race, and maternal education for the sample used in this report are summarized in Table 1.

**Table 1:**
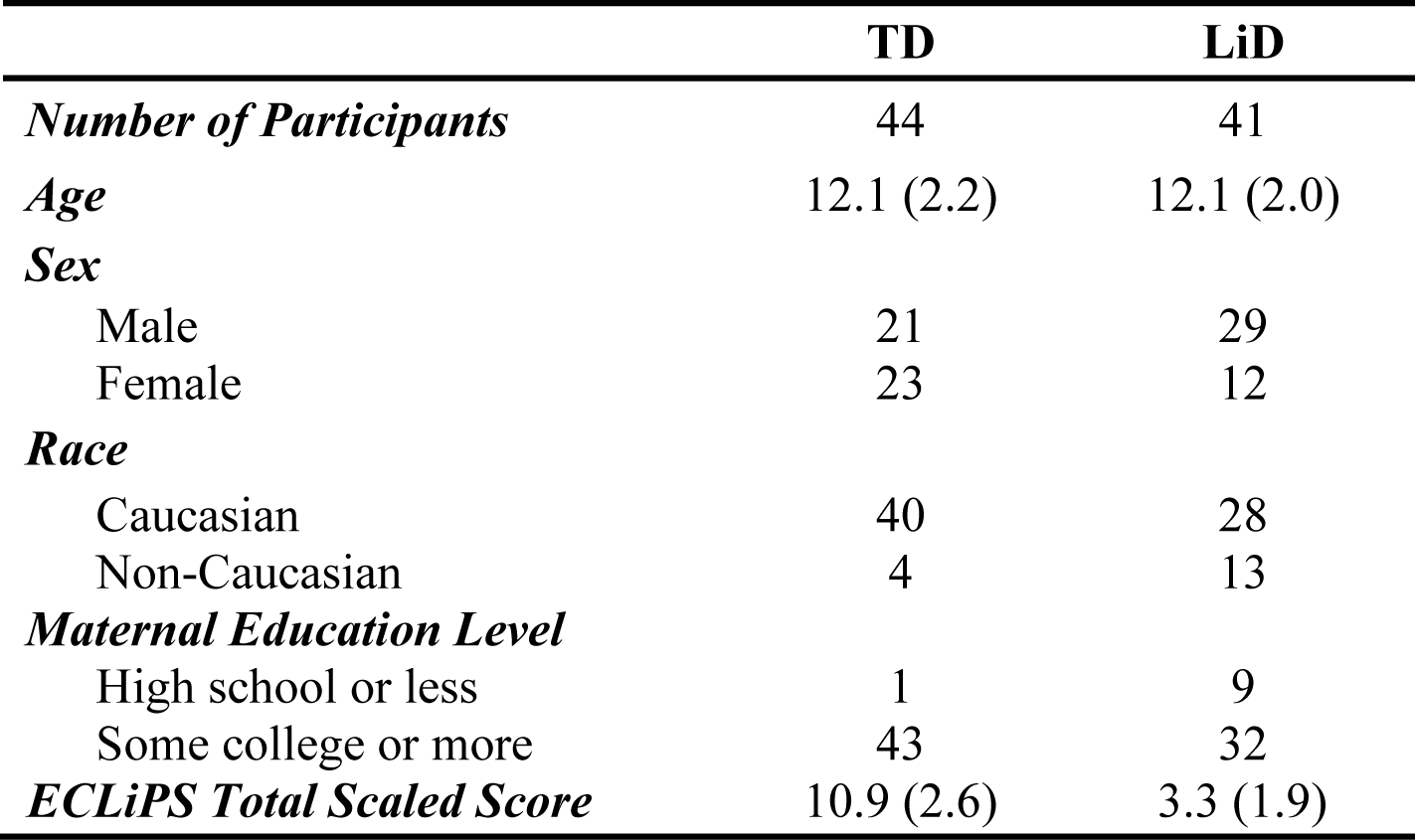
Demographics for the full study sample. Means and standard deviations are provided for age and the ECLiPS Total Scaled Score. All other values are frequencies.

|  | **TD** | **LiD** |
| --- | --- | --- |
| ***Number of Participants*** | 44 | 41 |
| ***Age*** | 12.1 (2.2) | 12.1 (2.0) |
| ***Sex*** |  |  |
| Male | 21 | 29 |
| Female | 23 | 12 |
| ***Race*** |  |  |
| Caucasian | 40 | 28 |
| Non-Caucasian | 4 | 13 |
| ***Maternal Education Level*** |  |  |
| High school or less | 1 | 9 |
| Some college or more | 43 | 32 |
| ***ECLiPS Total Scaled Score*** | 10.9 (2.6) | 3.3 (1.9) |

### Procedures

The overarching design of the research program was longitudinal, and several other reports have been published using sub-samples of its data to address various cross-sectional (Hunter et al., 2020, 2023; D. R. Moore et al., 2020; Petley et al., 2021; Stewart et al., 2022) and longitudinal (Kojima et al., in revision) questions regarding the nature of LiD. Participants in the research program completed a battery of behavioral tests for auditory and cognitive function, as well as neuroimaging using magnetic resonance imaging and EEG. The test battery for the present study, as well as the variables derived from it, is depicted in Figure 1. Due to differences in subject availability and data quality, as well as the requirements for some evoked responses, not all participants had the necessary data for all analyses.

**Figure 1.**
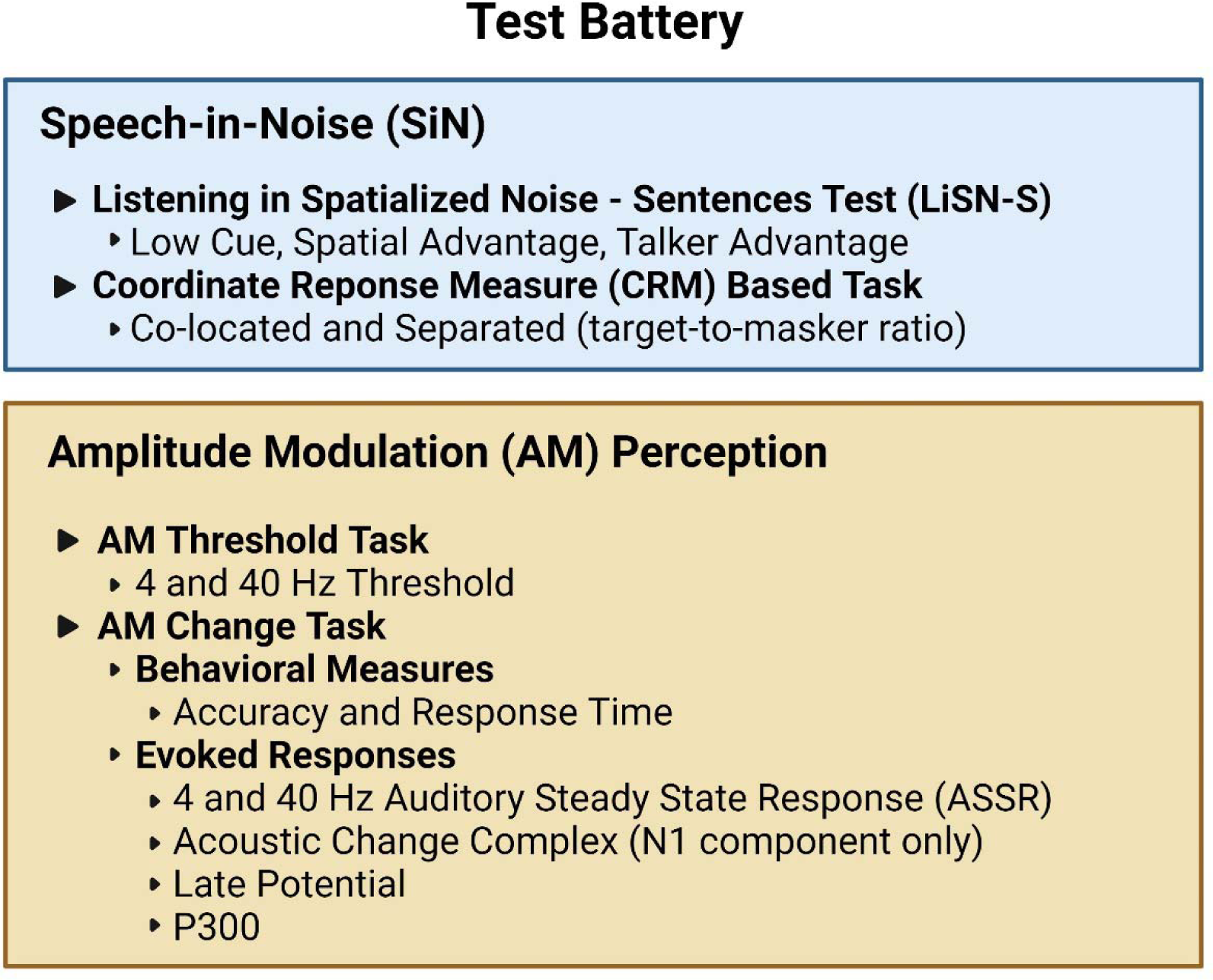
A summary of the study test battery and associated variables. Note that both evoked responses and behavioral measures were obtained during the AM Change task.

### Audiometry

Audiometric testing was completed for air conduction thresholds at standard octave test frequencies from 0.25 to 8 kHz as well as four extended high frequencies (10, 12.5, 14, and 16 kHz). Participants with elevated thresholds (> 20 dB HL) at the standard frequencies were excluded from the present analysis. As reported elsewhere, children with LiD did not differ from their TD peers on any measure of peripheral auditory function, including pure tone audiometry at standard and extended high frequencies, distortion product and chirp transient evoked otoacoustic emissions, middle ear reflexes, or wideband absorbance tympanometry (Hunter et al., 2020).

### Caregiver-Reported Listening Difficulties

Caregiver assessments of participants’ listening and communication abilities were collected via the ECLiPS questionnaire (Barry et al., 2015; Barry & Moore, 2014). The ECLiPS is composed of 38 items describing commonly observed behaviors related to listening and communication in children. The questionnaire asks caregivers to rate their degree of agreement with each statement on a five-point Likert scale. Responses on the ECLiPS can be summarized via five subscales, three composite scores, or a total score. These scores are age-scaled and standardized for a population mean of 10 (SD = 3) on the basis of British data (Barry et al., 2015). Inclusion in the LiD group was based on an ECLiPS total scaled score < 7 or a previous diagnosis of APD. Twelve children in the LiD group had a diagnosis of APD. ECLiPS total scaled scores for the LiD and TD samples used for the present report are provided in Table 1.

### Speech-in-Noise Tasks

The Listening in Spatialized Noise – Sentences test (LiSN-S; Brown et al., 2010; Cameron & Dillon, 2007; Phonak/NAL, 2011) permits the assessment of speech comprehension in the presence of informational masking. The U.S. edition of the task was administered using a commercial CD (Phonak/NAL, 2011) using a laptop computer with a task-specific soundcard and Sennheiser HD 215 headphones. The LiSN-S requires participants to repeat target sentences that are presented in the presence of speech from two distracting talkers. To evaluate the benefit obtained from different auditory cues, these distracting talkers vary with respect to their voice (same as the target or different) or their location (co-located with the target at 0° azimuth or separated at 90° azimuth while the target remains at 0°). Spatial locations are simulated via the use of generic head-related transfer functions of Humanski and Butler (1988). Three derived scores, called the Talker Advantage, Spatial Advantage, and Total Advantage scores, are obtained through subtraction processes between these conditions. The Talker Advantage reflects the improved speech reception threshold (SRT) when distracting talkers have a different voice than the target (co-located same voice vs. co-located different voice). The Spatial Advantage reflects the SRT improvement when the distracting talkers are spatially separated from the target (co-located same voice vs. separated same voice). The Total Advantage reflects the SRT reduction when both cues are available (co-located same voice vs. separated different voice). The rationale for these subtraction measures is that they isolate auditory processing from cognitive factors like selective attention (Moore & Dillon, 2018). The present analysis uses only the age-normed scores of the LiSN-S.

A task designed by Gallun and colleagues (2013) using stimuli from the Coordinate Response Measure (Bolia et al., 2000) was also used to measure auditory thresholds for target speech in the presence of speech maskers (Gallun et al., 2013). This task, referred to as the CRM-based task, was delivered via iPad (Apple Inc., Cupertino, CA) using Sennheiser HD 25 headphones while participants were seated in a sound-attenuating audiometric booth or quiet office. For every trial, the participant hears the phrase “Ready Charlie, go to (Color) (Number) now,” and is instructed to select the button with the spoken color and number on the iPad. Feedback is provided after every trial. The task includes three different conditions including a Single Talker (no masker; to ensure audibility of the target), Co-Located (target and masker presented at 0° azimuth) and Separated Talkers (target at 0° azimuth; maskers at ± 45° azimuth). Talker locations are simulated using generic head-related transfer functions. Performance on both masked conditions is expressed as a target-to-masker ratio (TMR).

### AM Perception Tasks

The AM perception tasks used in this study were modeled after the methods of Han and Dimitrijevic (2020), who employed two types of tasks: an AM Threshold task and an AM Change task (Han & Dimitrijevic, 2020). The AM Threshold task, which was used to measure behavioral AM detection thresholds, was conducted once for each modulation rate (4 and 40 Hz). The task was implemented as a custom MATLAB script and employed a three-interval forced choice task with trial-by-trial feedback. Each trial consisted of three consecutive 1-second segments of white noise, one of which contained AM. Participants were instructed to identify which stimulus contained AM and modulation depth was adaptively varied according to a 2 down, 1-up procedure with 2 dB steps. AM stimuli were level-matched to non-modulated noise segments via root mean square matching. The task terminated after nine reversals, and the resulting thresholds were the average of the last six.

The primary purpose of the AM Change task was to obtain evoked responses to changes in AM rate (i.e., the ACC and P300), but it also yielded behavioral performance metrics (accuracy and response time, RT). The task involved listening to continuous white noise that contained AM. The parameters of these stimuli were identical to the AM Threshold task, except that the noise was delivered continuously with 100% modulation depth. Alternations between the two AM rates (4 and 40 Hz) occurred at random intervals between 2 and 3 seconds. Participants were asked to listen continuously for these changes and press a response button as quickly and accurately as possible if they detected one. Stimuli for both AM tasks were presented via ER-3 insert earphones, to the right ear only at 72 dBA for the AM Threshold task and 70 dBA for the AM Change task. All tasks were carried out with participants seated in a Faraday-shielded double-walled sound booth.

### Electroencephalography

The EEG data was collected continuously during the AM Change task using a 64-channel actiCHamp system (Brain Products, GmbH, Inc., Munich, Germany). Electrodes were mounted in an elasticized cap with an equidistant layout arranged around a vertex sensor located at Cz of the 10-20 system. Some participants were additionally fitted with single electrodes below the right eye, on each mastoid, and on the tip of the nose (Nz). The EEG data was collected at 2000 Hz and stored for offline analysis. Some participant data was collected with Cz as the online reference, while others were referenced to Nz.

### Data Analysis

#### AM Change Task Performance

Responses on the AM Change task were deemed correct if they occurred between 100 and 2135 ms from change onset. These limits represented the approximate minimum RT for voluntary responses to auditory stimuli (Pain & Hibbs, 2007; Thompson et al., 1992) and the threshold for extreme outlier RTs for LiD subjects on this task when all responses were accepted. Accurate RT information could not be obtained for 17 participants; thus, they were excluded from all analyses of behavioral performance on the AM Change task.

#### Evoked Responses

Analyses of the EEG data were carried out using Matlab R2018b (Mathworks, Inc.), via a combination of custom scripts, EEGLAB v13.6.5b (Delorme & Makeig, 2004), and ERPLAB v8.0 (Lopez-Calderon & Luck, 2014). The data was visually inspected for channels with poor data quality and segments of data that violated the assumption of stationarity for decomposition via independent component analysis (ICA) with respect to either amplitude (> 100 µV) or frequency (e.g., sporadic muscle artifacts). Such segments and channels were rejected prior to ICA decomposition. Independent components were extracted using Infomax ICA (as implemented in EEGLAB) on data that was re-referenced to the average reference and band-pass filtered between 2 and 30 Hz using a 2^nd^ order Butterworth filter applied in the forward and backward directions (Klug & Gramann, 2021; Winkler et al., 2015). These components were used to correct continuous data which was re-referenced to the average reference and filtered using 2^nd^ order Butterworth filters using the following bandpass settings for each evoked response: 0.1 – 30 Hz for cortical ERPs, 0.5 – 20 Hz for the 4 Hz ASSR, and 20 – 60 Hz for the 40 Hz ASSR. Note that this re-referencing process is algebraically identical to collecting all datasets with the same reference and entirely eliminates any possible differences due to the choice of online reference electrode. Only components that were deemed to reflect ocular or cardiac sources were removed. Electrodes that were previously removed due to poor data quality were then interpolated when possible. *ASSR.* To compute the ASSR, data from homogeneous periods of AM stimulation was segmented into 1-second epochs and any epochs whose maximum absolute voltage was in the top 15% of all values were automatically identified and rejected from further analyses. The ASSR was quantified using SNR, a measure of ASSR signal (*s*) amplitude versus other neural “noise” (*n*), and phase coherence. Phase coherence indexes the replicability of the response latency across epochs. It was quantified in sweeps that consisted of sixteen 1.024-second epochs according to the methods of Picton and colleagues (2001) using normal (i.e., non-weighted) averaging. The power (*P*) of the neural background noise (*n*) was estimated from the 60 neighboring frequency bins on either side of the response frequency bins and the SNR of the ASSR was calculated as:

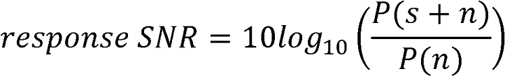

The significance of the ASSR at each electrode was determined using an *F*-statistic derived by assessing this SNR against the *F* distribution with 2 and 240 degrees of freedom (Picton et al., 2001). This procedure yields significance with an SNR of 4.8 dB. All negative SNRs were changed to a baseline of 0 dB. Phase coherence (*R*) is a value between 0 and 1, with larger values corresponding to a lower probability that the phase of the response (*θ*) is changing randomly between epochs (Picton et al., 2001), and was calculated as:

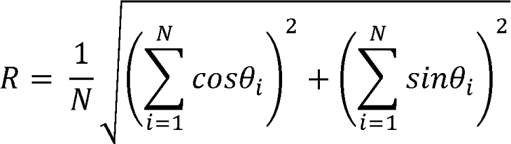

To provide balanced comparisons between participants, all metrics were reported from the average corresponding to the maximum number of sweeps (*N*) that was available across all participants (22 sweeps). This is well within the range of sweeps used for measurement of the ASSR in pediatric samples, which varies considerably between studies (e.g., De Vos et al., 2020; Swanepoel et al., 2004; Vanvooren et al., 2014, 2015). Significant ASSRs were observed at both frequencies (4 and 40 Hz) for all participants, across many electrodes (4 Hz *M* = 28.0 electrodes, *SD* = 7.7; 40 Hz *M* = 35.8 electrodes, *SD* = 3.8). Similarly to other research examining the ASSR and its lateralization in pediatric samples, the ASSR was quantified separately over the left and right hemispheres in a subset of parieto-occipital electrodes (Left hemisphere: E6, E7, E15, E16, E17, E18, E20, Right hemisphere: E24, E25, E33, E34, E36, E37, E39), and left-handed participants were excluded (De Vos et al., 2020; Vanvooren et al., 2014, 2015). Handedness was determined based on caregiver report.

##### Cortical event-related potentials (ERPs)

For the analyses of the ACC, a late potential following the ACC (called the LP), and P300, the continuous data was segmented into 900 ms epochs extending from 100 ms pre-AM rate change to 800 ms post-AM rate change for target events (referred to as “Change” epochs) and encompassing 900 ms of homogeneous AM for epochs containing no target event (referred to as “No Change” epochs). Though the ACC correlates well with perceptual discrimination thresholds, it does not require attention and is commonly measured under conditions of passive stimulation (e.g., Han & Dimitrijevic, 2015, 2020; He et al., 2012; Martin & Boothroyd, 2000; Uhler et al., 2018). Thus, it was computed using all “Change” epochs, regardless of whether the target was accurately detected. Artifact-contaminated epochs were automatically identified using an absolute voltage threshold of +/- 75 µV and a peak-to-peak voltage threshold of 100 µV. Some rare participants (3 TD and 1 LiD subject for the ACC, and 1 LiD subject for the P300) demonstrated high amplitude alpha that prevented the use of these thresholds. In these cases, a more liberal absolute voltage threshold of +/- 95 µV was applied, with no peak-to-peak threshold.

The N1 of the ACC was measured in difference waves computed by subtracting the averaged no change waveform from the target waveform, regardless of AM rate or direction of change. The amplitude of N1 was measured at the frontocentral midline site anterior to Cz (electrode e2) as a mean amplitude in the +/- 20 ms window surrounding its visually identified peak in the grand average waveforms for target epochs. The peak was identified separately for the LiD and TD groups. Owing to the active nature of the task, the P2 of the ACC was not discernable as a separate peak from P300, which itself was complex and multi-peaked. A LP, likely reflecting contributions from both P2 and P300, was measured in the same waveforms as the ACC as an area under the curve at the centroparietal midline site directly posterior to Cz (e35) for all positive peaks starting at its visually identified onset in the grand difference waveforms. As with the N1, the latency for the onset of the LP was identified separately for the LiD and TD groups.

Unlike the ACC, the P300 is only observed when selective attention is actively directed towards the task stimuli. As outlined by Duncan and colleagues (2009) in their formative review on best practices in the measurement of cognitive ERPs, the P300 should be measured from an average of at least 36 correctly-detected targets. That approach was used here. Similarly to the LP, P300 was measured as an area under the curve directly posterior to Cz (e35) for all positive peaks starting at its visually identified onset in the grand difference waveforms, and its onset was evaluated separately for the LiD and TD groups. Data quality metrics including the number of epochs accepted for averaging, the ERP measurement window, and the number of ICA components rejected per participant are summarized in Supplementary Table 1.

#### Statistical Analyses

Correlational analyses were carried out to (1) explore the effect of age on AM and SiN perception, (2) examine the relationships between measures of AM and SiN perception, and (3) quantify the relationships between behavioral and evoked response measures of AM perception. Since study procedures were completed at different times for different participants, the analysis of age required the exclusion of any participants whose age differed substantially (> 3 months) between any two variables of interest. Since the scores from the LiSN-S were age-normed, all correlations that were computed against them employed variables that were age-adjusted via linear regression using the study sample. All correlations were computed as Spearman rank order coefficients across all participants, regardless of group. This approach maximized statistical power and permitted the examination of these relationships across a broader range of auditory skills than would be available in a purely TD population. Secondary correlations were also computed within each group, but since these analyses were under-powered, they were not interpreted for their statistical significance. Rather, they served as effect size estimates to help interpret the magnitude of the observed relationships as a function of LiD.

To examine the basis of LiD, behavioral and evoked response measures of AM perception were compared between children with LiD and their TD peers using age-matched groups. Since EEG data collection did not occur for all participants, and technical challenges led to some loss of some response data, the sample sizes for these comparisons differed. Behavioral performance and cortical ERPs from the AM Change task were compared between the two groups via *t*-tests when the assumption of normality was met and Wilcoxon rank-sum tests when it was not. Effect sizes were computed using Cohen’s *d*.

Analyses for the ASSR and AM Thresholds were carried out via mixed-design ANOVAs. For the ASSR, two three-way ANOVAs were performed, one for SNR and one for phase coherence, both including group, hemisphere, and modulation rate (4 vs. 40 Hz) as factors. Effect sizes for these ANOVAs were reported as generalized eta squared (η^2^_G_, Olejnik & Algina, 2003). For the AM thresholds, which tended not to be normally distributed, a nonparametric (aligned ranks) ANOVA was carried out with the factors group and modulation rate. Effect sizes were reported as partial eta squared (η_p_^2^, Cohen, 1973). Posthoc analyses to explore interactions following this ANOVA were carried out using Wilcoxon rank sum tests with Holm-Bonferroni correction.

To counteract alpha inflation without being overly conservative, all correlational and paired contrast *p*-values were adjusted using Bonferroni correction in a familywise manner (Rubin, 2017). These families were constructed based on the number of metrics that were derived from the same test or based on known correlations between independent tests. Thus, for tests involving any of the three measures derived from the LiSN-S, *p*-values were corrected for a family size of three. Similarly, for tests using the CRM-based task, which yielded two measures, *p*-values were corrected for two comparisons. Since accuracies and RTs tend to correlate with one another (Draheim et al., 2021), any tests involving them were also corrected for two comparisons. For the ASSR, since both SNR and phase coherence were measured, tests involving them were corrected for a family size of two. Finally, since there was an overlap in the data used for the LP and P300, tests involving them were corrected for two comparisons. In cases where more than one family was involved in a statistical test, these correction factors were multiplied. Thus, for example, when correlations were computed between measures from the LiSN-S (family size = 3) and accuracy on the AM change task (family size = 2), the correction factor was six.

## Results

### Relationships Between Measures of AM and SiN Perception

#### Age Effects

Correlations between age and all variables of interest are summarized in Table 2. These tended to be low and very few variables exhibited significant relationships. Among the behavioral measures of AM perception, only performance on the AM Threshold task at 4 Hz correlated significantly with age, and indicated that older children had lower (i.e., better) thresholds. The observation of similar effect sizes within each of the groups suggests that this relationship is not affected by LiD. While age had no significant relationship with accuracy on the AM Change task in the across-group analysis, it remains possible that accuracy increases with age for children with LiD given the medium effect size that was observed in this group, *r_s_*(12) = 0.60, but not their TD peers, *r_s_*(20) = 0.07. Consistent with the fact that LiSN-S scores were age-normed, among the SiN tasks, only the CRM-based task exhibited any correlations with age, specifically in the Separated Talker condition. These correlations indicated that older children performed better (i.e., had lower TMRs) in the Separated Talker condition of the CRM-based task than younger children. This effect may have been stronger in TD children, *r_s_*(43) = −0.55, than in those with LiD, *r_s_*(37) = −0.20. Scatterplots illustrating correlations between behavioral metrics of AM perception or SiN and age are shown in panel A of Figure 2 for all relationships that reached significance across the two groups.

**Figure 2.**
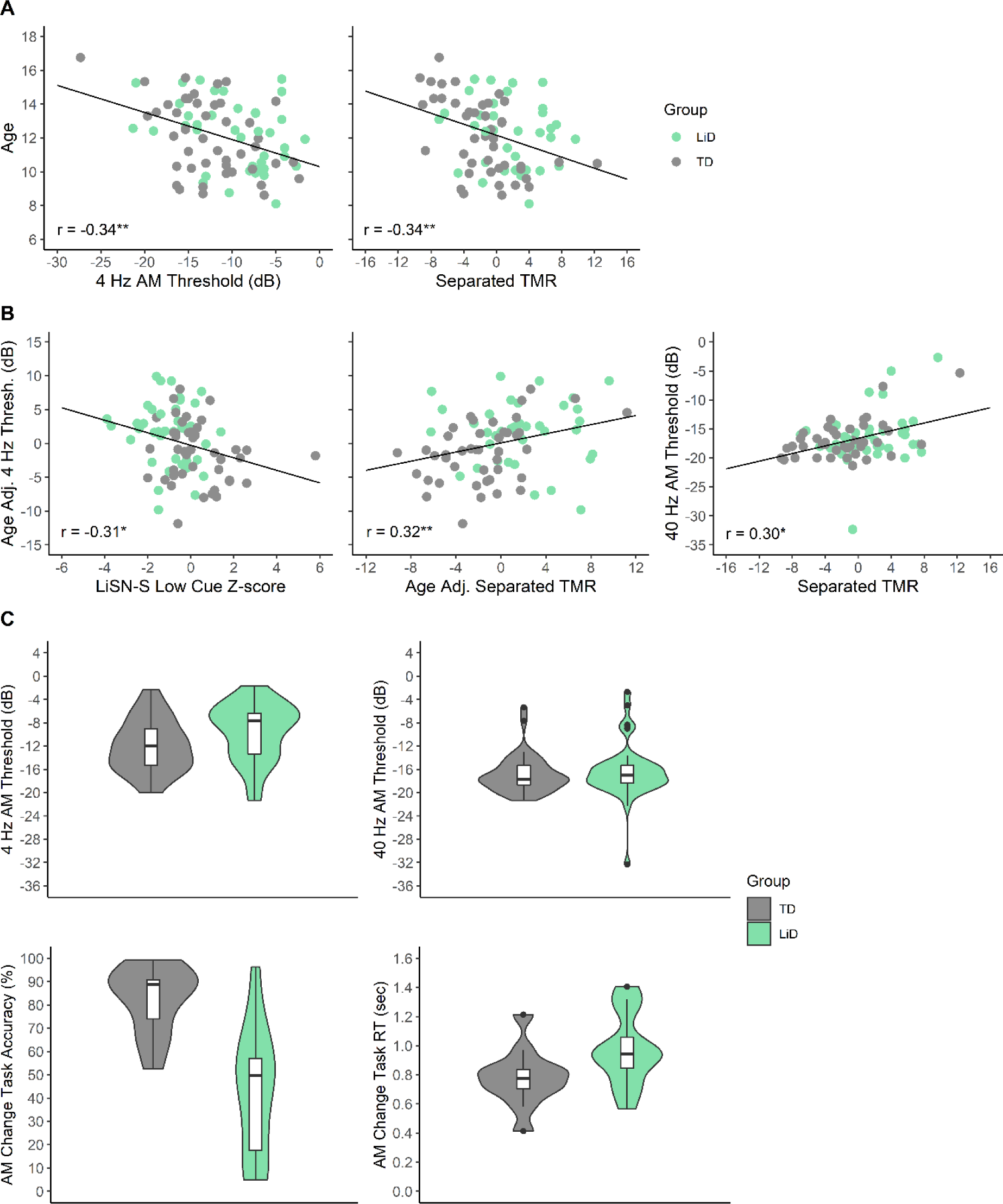
**A and B:** Scatterplots illustrating the significant correlations between (A) age and behavioral measures of AM and SiN perception, and (B) behavioral measures of AM perception and SiN performance. Separated TMR = Separated Talker target-to-masker ratio of the CRM-Based Task, \**p* < 0.05, \*\**p* < 0.01. While scores from the LiSN-S were age-corrected based on test norms, other scores were age-adjusted based on the present sample, where specified. Correlations were computed using all participants’ data, regardless of group. **C:** Group scores on behavioral measures of AM perception. Superior performance for the TD group was evident for the 4 Hz AM threshold and accuracy on the AM Change task.

**Table 2:** Spearman correlations between age and metrics from the AM perception and SiN tasks. Owing to the low statistical power of the within-group analyses, these coefficients are interpreted as effect sizes only and *p*-values are not provided.

|  | Across Groups |  |  | TD |  | LiD |  |
| --- | --- | --- | --- | --- | --- | --- | --- |
| | N | $r_s$ | $p$ | N | $r_s$ | N | $r_s$ |
| ***LiSN-S*** |  |  |  |  |  |  |  |
| Low Cue | 84 | -0.09 | 1.00<sup>†</sup> | 44 | 0.08 | 40 | -0.28 |
| Spatial Advantage | 84 | -0.10 | 1.00<sup>†</sup> | 44 | -0.16 | 40 | -0.01 |
| Talker Advantage | 84 | -0.14 | .568<sup>†</sup> | 44 | -0.22 | 40 | -0.05 |
| ***CRM-based Task*** |  |  |  |  |  |  |  |
| Co-located | 80 | -0.04 | 1.00<sup>†</sup> | 43 | 0.09 | 37 | -0.09 |
| Separated | 80 | -0.34 | .004<sup>†</sup> | 43 | -0.55 | 37 | -0.20 |
| ***AM Threshold*** |  |  |  |  |  |  |  |
| 4 Hz | 85 | -0.34 | .001 | 44 | -0.36 | 41 | -0.33 |
| 40 Hz | 85 | -0.09 | .411 | 44 | -0.19 | 41 | < 0.01 |
| ***AM Change Task*** |  |  |  |  |  |  |  |
| Accuracy | 32 | 0.20 | .565<sup>†</sup> | 20 | 0.07 | 12 | 0.60 |
| Response Time | 32 | -0.31 | .377<sup>†</sup> | 20 | -0.31 | 12 | -0.15 |
| ***ASSR SNR*** |  |  |  |  |  |  |  |
| 4 Hz | 40 | -0.10 | 1.00<sup>†</sup> | 26 | -0.20 | 14 | 0.23 |
| 40 Hz | 40 | 0.30 | .114<sup>†</sup> | 26 | 0.24 | 14 | 0.36 |
| ***ASSR Phase Coherence*** |  |  |  |  |  |  |  |
| 4 Hz | 40 | -0.10 | 1.00<sup>†</sup> | 26 | -0.22 | 14 | 0.23 |
| 40 Hz | 40 | 0.22 | .342<sup>†</sup> | 26 | 0.15 | 14 | 0.31 |
| ***Cortical ERPs*** |  |  |  |  |  |  |  |
| N1 Amplitude | 48 | -0.31 | .031 | 30 | -0.31 | 18 | -0.29 |
| LP Area | 48 | 0.01 | 1.00<sup>†</sup> | 30 | 0.17 | 18 | -0.48 |
| P300 Area | 27 | 0.06 | 1.00<sup>†</sup> | 18 | 0.25 | 9 | -0.52 |

<sup>†</sup>Bonferroni-corrected

Among the evoked responses, only the amplitude of the N1 exhibited a significant correlation with age, with older children showing larger (i.e., more negative) N1 responses. This effect was similar across the two groups [LiD *r_s_*(18) = −0.29, TD *r_s_*(30) = −0.31], and is illustrated in panel A of Figure 3. Unlike N1, there was no significant effect of age on the areas of the LP or P300 across the two groups, though the coefficients obtained separately for the groups suggest that the P300 may vary with age for children with LiD, *r_s_*(9) = −0.52.

**Figure 3.**
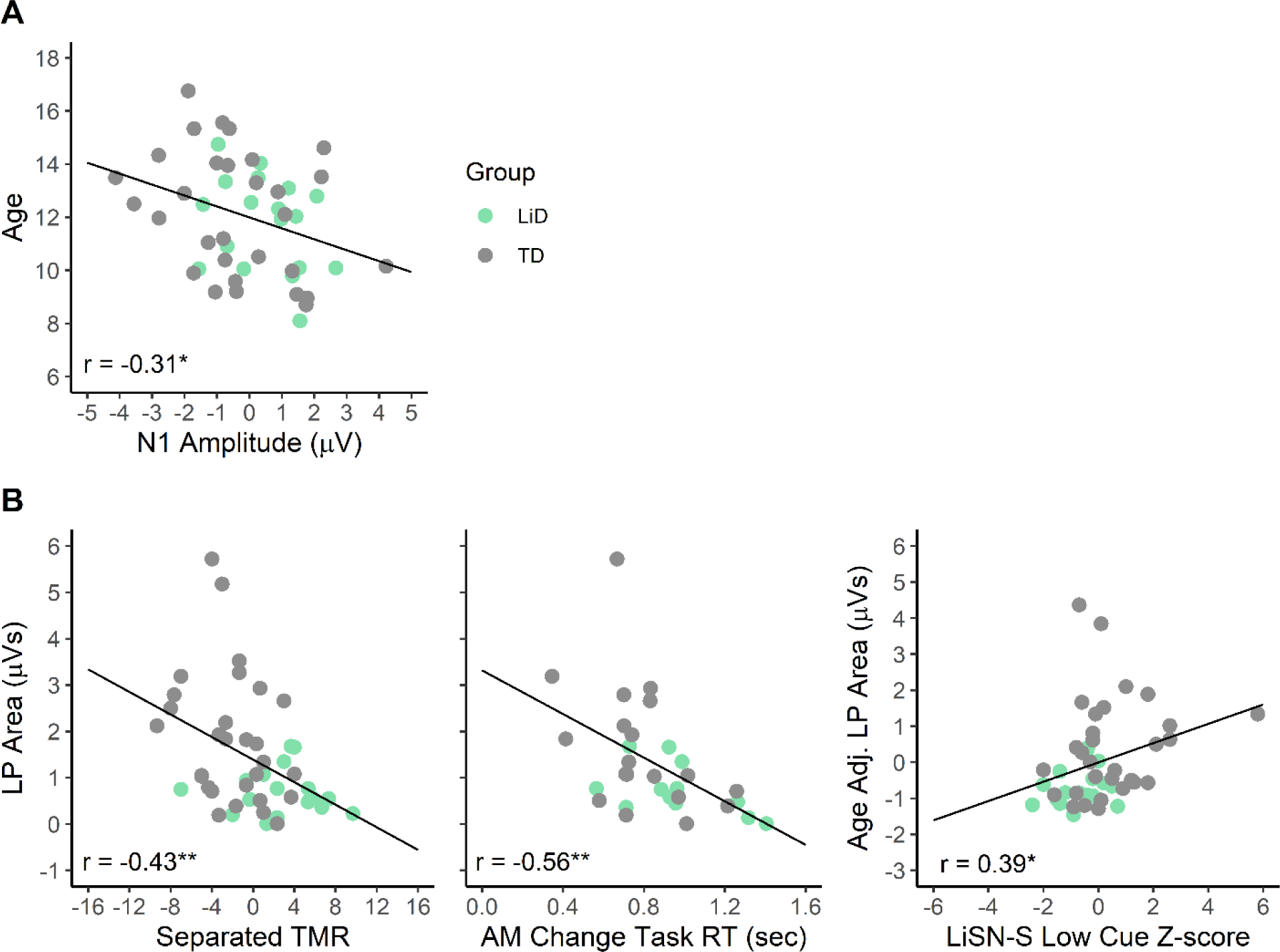
Scatterplots for all significant correlations between cortical ERPs and (A) age as well as (B) behavioral metrics of AM and SiN perception. Note that correlations were computed across the two groups, with LiSN-S scores age-adjusted based on test norms and other scores age-adjusted based on the present sample, where specified. Separated TMR = Separated Talker target-to-masker ratio of the CRM-Based Task, LP = late potential, \**p* < 0.05, \*\**p* < 0.01

#### SiN Correlations

The relationships between age-adjusted measures from the AM perception tasks and performance on the LiSN-S are summarized in Table 3. Very few significant correlations were observed. Specifically, 4 Hz AM thresholds and the area under the LP curve correlated significantly with the Low Cue condition of the LiSN-S. These correlations indicated that children with lower (poorer) scores in the Low Cue condition of the LiSN-S had higher (poorer) thresholds for detecting 4 Hz AM and smaller LPs. Scatterplots illustrating these relationships are shown in panel B of Figures 2 and 3, respectively. Within-group correlations suggest that the relationship between the 4 Hz AM threshold and performance in the Low Cue condition of the LiSN-S was similar between the two groups [TD *r_s_*(44) = −0.25, LiD *r_s_*(40) = - 0.18], as was the relationship with the area under the LP curve [TD *r_s_*(30) = 0.23, LiD *r_s_*(18) = 0.24]. Within-group correlations yielded a medium effect size, *r_s_*(18)= −0.58, for the relationship between LiSN-S Spatial Advantage scores and the amplitude of the N1 component, for the LiD group only. The effect did not reach significance in the across-group analysis.

**Table 3:** Spearman correlations between age-corrected measures from the AM perception tasks and age-normed scores from the LiSN-S. The LiSN-S Low Cue score is obtained with co-located target and masker speech spoken by the same talker. The Spatial and Talker Advantage scores are derived by comparing this condition against those with spatial separation or a change in voice for the masker speech. All *p*-values were Bonferroni corrected for multiple comparisons. Within-group correlation coefficients are interpreted as effect sizes only.

|  | Across Groups |  |  |  | TD |  |  |  | LiD |  |  |  |
| --- | --- | --- | --- | --- | --- | --- | --- | --- | --- | --- | --- | --- |
|  | N | Low Cue | Spat. Adv. | Talk. Adv. | N | Low Cue | Spat. Adv. | Talk. Adv. | N | Low Cue | Spat. Adv. | Talk. Adv. |
| ***AM Threshold*** |  |  |  |  |  |  |  |  |  |  |  |  |
| 4 Hz | 84 | -0.31* | -0.03 | -0.17 | 44 | -0.25 | 0.02 | 0.03 | 40 | -0.18 | -0.07 | -0.28 |
| 40 Hz | 84 | -0.22 | -0.05 | -0.20 | 44 | -0.21 | 0.03 | -0.04 | 40 | -0.17 | -0.13 | -0.34 |
| ***AM Change Task*** |  |  |  |  |  |  |  |  |  |  |  |  |
| Accuracy | 32 | -0.07 | 0.10 | 0.12 | 20 | -0.43 | 0.30 | 0.14 | 12 | 0.07 | -0.33 | -0.14 |
| Response Time | 32 | 0.25 | -0.21 | -0.02 | 20 | 0.64 | -0.26 | 0.26 | 12 | -0.12 | -0.04 | -0.31 |
| ***ASSR SNR*** |  |  |  |  |  |  |  |  |  |  |  |  |
| 4 Hz | 40 | 0.01 | 0.22 | 0.01 | 26 | 0.04 | 0.27 | < 0.01 | 14 | 0.05 | 0.08 | 0.33 |
| 40 Hz | 40 | -0.15 | 0.16 | 0.09 | 26 | -0.14 | 0.22 | 0.02 | 14 | -0.24 | -0.03 | 0.17 |
| ***ASSR Phase Coherence*** |  |  |  |  |  |  |  |  |  |  |  |  |
| 4 Hz | 40 | 0.08 | 0.21 | 0.01 | 26 | 0.12 | 0.24 | -0.03 | 14 | 0.07 | 0.07 | 0.31 |
| 40 Hz | 40 | -0.13 | 0.20 | 0.14 | 26 | -0.16 | 0.22 | 0.02 | 14 | -0.16 | 0.21 | 0.35 |
| ***Cortical ERPs*** |  |  |  |  |  |  |  |  |  |  |  |  |
| N1 Amplitude | 48 | -0.08 | -0.23 | -0.07 | 30 | 0.09 | -0.15 | 0.09 | 18 | -0.11 | -0.58 | -0.22 |
| LP Area | 48 | 0.39* | 0.01 | -0.09 | 30 | 0.23 | -0.16 | -0.29 | 18 | 0.24 | 0.37 | 0.08 |
| P300 Area | 27 | 0.21 | 0.17 | -0.09 | 18 | 0.15 | -0.02 | -0.19 | 9 | 0.40 | 0.52 | -0.06 |

\* $p < 0.05$

Correlations between measures from the AM perception tasks and performance on the CRM-based task are summarized in Table 4. Significant relationships were only observed for the Separated Talker condition. Similarly to the Low Cue condition of the LiSN-S, this condition had a significant relationship with AM thresholds, but in this case, both modulation rates were implicated. These correlations, illustrated in panel B of Figure 2, indicated that children who had higher (poorer) thresholds for detecting AM also exhibited higher TMRs (poorer performance) on the CRM-based task. Since scores in the Separated Talker condition of the CRM-based task and AM Thresholds at 4 Hz were both correlated with age, a second correlation was computed using the residuals of linear models with age as the predictor. These factors continued to be significantly correlated following the removal of age-related variance, *r_s_*(84) = 0.32, adj. *p* = .007, indicating that the relationship between the unadjusted Separated Talker condition of the CRM-based task and 4 Hz AM Thresholds cannot be attributed entirely to maturation. Within-group effect sizes suggested that the across-group correlation between the Separated Talker condition and 4 Hz AM thresholds may have been driven primarily by the TD group, even after age-adjusting both variables [TD *r_s_*(43) = 0.40, LiD *r_s_*(37) = 0.04]. By contrast, there was little difference in effect size between the groups [TD *r_s_*(43) = 0.34, LiD group *r_s_*(37) = 0.23] for the relationship with 40 Hz thresholds.

**Table 4:**
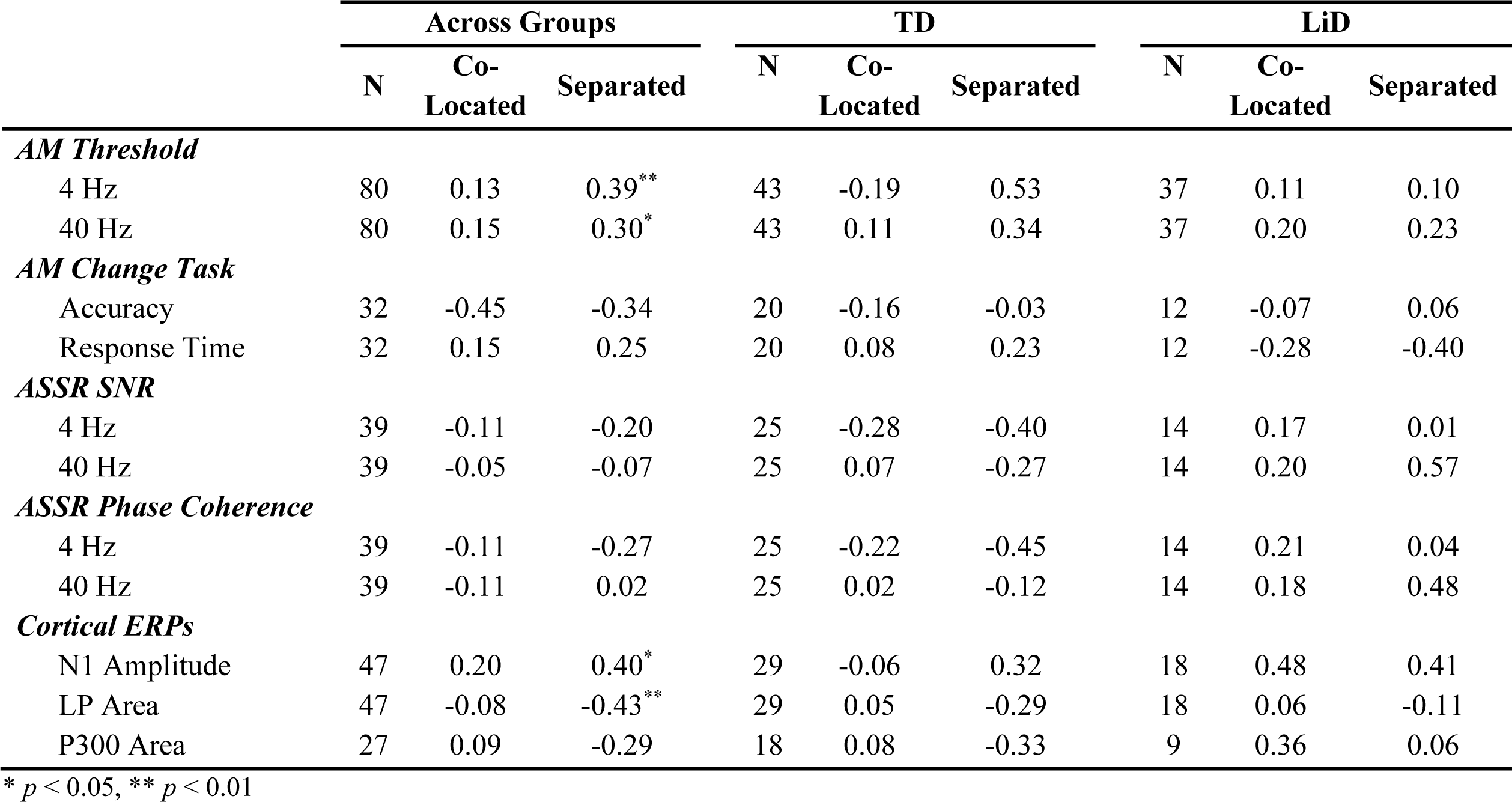
Spearman correlations between measures from the AM perception tasks and performance on the CRM-based task. All *p*-values have been Bonferroni corrected for multiple comparisons. Within-group coefficients are interpreted as effect sizes only.

The across-group analysis also yielded significant relationships between performance in the Separated Talker condition of the CRM-based task and two evoked responses: the amplitude of the N1 and the area under the LP curve. These correlations indicated that children with larger LPs and N1 amplitudes had lower (better) TMRs. The correlation involving the area under the LP curve may have been driven by the TD group [*r_s_*(29) = −0.29] somewhat more than the LiD group [*r_s_*(18) = −0.11]. A scatterplot illustrating this relationship is shown in Panel B of Figure 3. Due to their common relationships with age, a second correlation was computed between the Separated Talker TMR and the amplitude of the N1 following the removal of variance associated with age. In this correlation, the relationship between N1 amplitude and Separated Talker TMRs lost significance, *r_s_*(47) = 0.32, adj. *p* = .056. The LiD group also exhibited a moderate effect size [*r_s_*(14) = 0.57] for a relationship between the SNR of the 40 Hz ASSR and performance in the Separated Talker condition of the CRM-based task, but this correlation did not reach significance in the across-group analysis.

### Behavioral Versus Evoked Response Measures of AM Perception

The relationships between behavioral and evoked response measures derived from the AM perception tasks are summarized in Tables 5 and 6. Several reached statistical significance, the majority involving the 40 Hz ASSR. Higher SNRs and phase coherence for the 40 Hz ASSR were associated with lower (better) 4 Hz AM thresholds, and both of these relationships tended to hold true when measured separately for the two groups. Both the SNR and phase coherence of the 40 Hz ASSR also exhibited significant correlations with performance on the AM Change task, such that children with larger and more synchronized 40 Hz ASSRs achieved higher accuracy and responded faster when detecting changes in AM rate. These relationships are illustrated in Panel A of Figure 4. While the relationship between the 40 Hz ASSR and RT was fairly stable across groups, with respect to both SNR [TD *r_s_*(17) = −0.38, LiD *r_s_*(9) = −0.43] and phase coherence [TD *r_s_*(17) = −0.35, LiD *r_s_*(9) = −0.37], this was not the case for accuracy. Instead, correlations with accuracy tended to be driven by the TD group for both the SNR [TD *r_s_*(17) = 0.49, LiD *r_s_*(9) = 0.18] and phase coherence [TD *r_s_*(17) = 0.60, LiD *r_s_*(9) = 0.10]. As illustrated in Panel B of Figure 3, the area under the LP curve also correlated significantly with RT on the AM Change task, indicating that children with larger LPs had shorter RTs. Within-group correlations demonstrated that this relationship was present for both TD children [*r_s_*(19) = −0.51] and those with LiD [*r_s_*(12) = −0.47]. The LiD group also exhibited a medium-large effect size for a relationship between the area of the P300 and accuracy [*r_s_*(9) = −0.73], despite the lack of a significant correlation in the across-group analysis.

**Figure 4.**
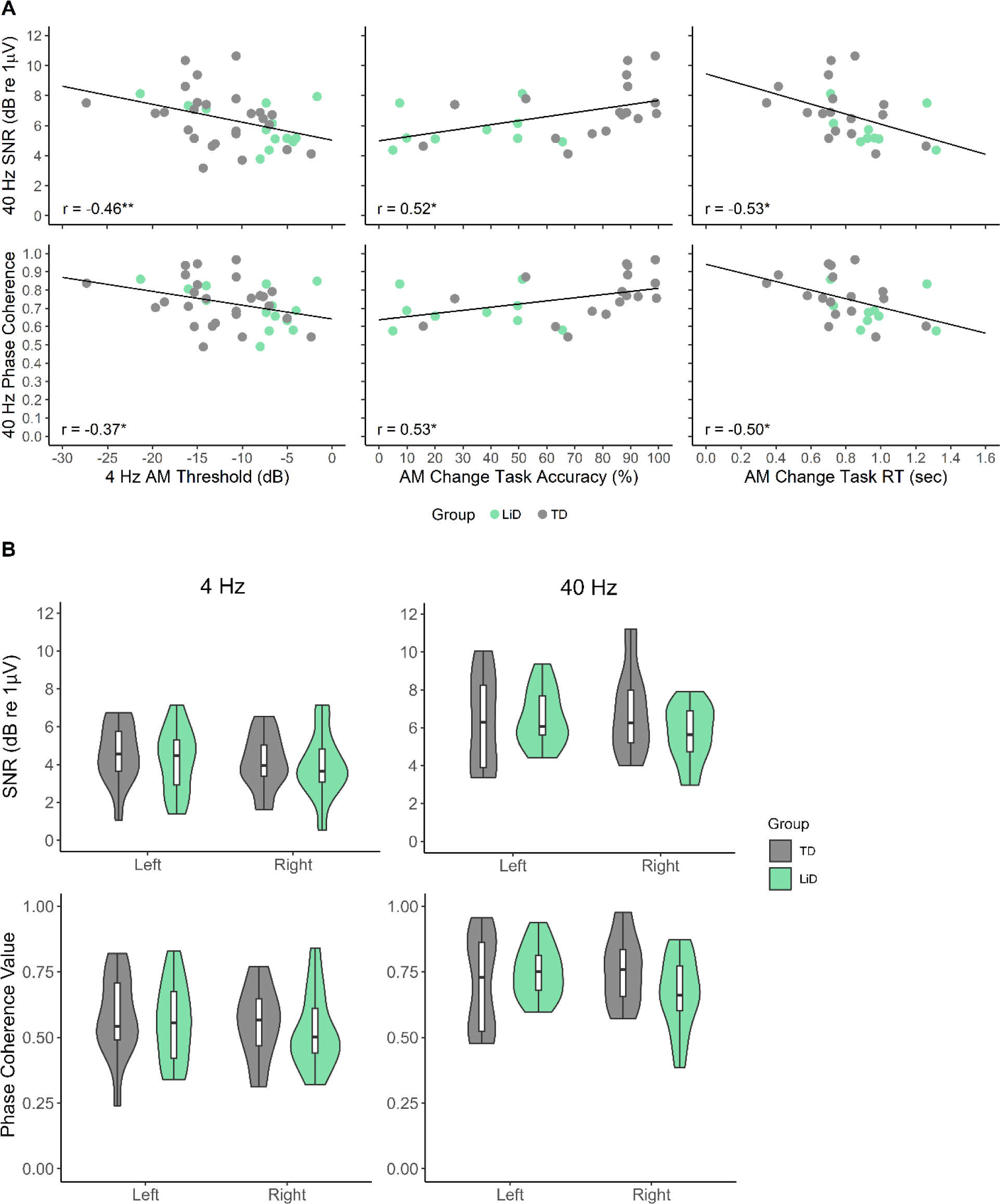
**A:** Scatterplots illustrating all significant correlations between ASSR metrics and behavioral measures of AM perception, \**p* < 0.05, \*\**p* < 0.01. All ASSR metrics were measured using a pool of parieto-occipital electrodes and correlations were computed across the two groups. **B:** ASSR metrics as a function of group, frequency, and hemisphere. A significant effect was only observed for frequency, in which both SNR and phase coherence were higher at 40 Hz.

**Table 5:** Spearman correlations between evoked responses from the AM Change task and AM detection thresholds. *p*-values have been adjusted for multiple comparisons where applicable. Within-group coefficients are interpreted as effect sizes only.

|  | Across Groups |  |  | TD |  |  | LiD |  |  |
| --- | --- | --- | --- | --- | --- | --- | --- | --- | --- |
|  | N | 4 Hz | 40 Hz | N | 4 Hz | 40 Hz | N | 4 Hz | 40 Hz |
| ***ASSR SNR*** |  |  |  |  |  |  |  |  |  |
| 4 Hz | 40 | -0.21 | -0.06 | 26 | -0.30 | -0.20 | 14 | -0.31 | 0.11 |
| 40 Hz | 40 | -0.46** | -0.17 | 26 | -0.43 | -0.32 | 14 | -0.34 | 0.28 |
| ***ASSR Phase Coherence*** |  |  |  |  |  |  |  |  |  |
| 4 Hz | 40 | -0.22 | -0.07 | 26 | -0.32 | -0.19 | 14 | -0.27 | 0.16 |
| 40 Hz | 40 | -0.37* | -0.13 | 26 | -0.24 | -0.21 | 14 | -0.31 | 0.19 |
| ***Cortical ERPs*** |  |  |  |  |  |  |  |  |  |
| N1 Amplitude | 48 | 0.13 | 0.13 | 30 | 0.09 | -0.03 | 18 | 0.14 | 0.30 |
| LP Area | 48 | -0.31 | -0.16 | 30 | -0.25 | -0.03 | 18 | 0.12 | -0.18 |
| P300 Area | 27 | -0.28 | -0.07 | 18 | -0.18 | 0.06 | 9 | -0.23 | -0.07 |

\* $p < 0.05$ , \*\* $p < 0.01$

**Table 6:** Spearman correlations between neural and behavioral measures from the AM Change task (RT = response time). *p*-values have been adjusted for multiple comparisons where applicable. Within-group analyses, these coefficients are interpreted as effect sizes only.

|  | Across Groups |  |  | TD |  |  | LiD |  |  |
| --- | --- | --- | --- | --- | --- | --- | --- | --- | --- |
|  | N | RT | Accuracy | N | RT | Accuracy | N | RT | Accuracy |
| ***ASSR SNR*** |  |  |  |  |  |  |  |  |  |
| 4 Hz | 26 | 0.01 | 0.01 | 17 | -0.01 | 0.24 | 9 | -0.32 | 0.33 |
| 40 Hz | 26 | -0.53* | 0.52* | 17 | -0.38 | 0.49 | 9 | -0.43 | 0.18 |
| ***ASSR Phase Coherence*** |  |  |  |  |  |  |  |  |  |
| 4 Hz | 26 | -0.02 | 0.01 | 17 | -0.04 | 0.22 | 9 | -0.32 | 0.33 |
| 40 Hz | 26 | -0.50* | 0.53* | 17 | -0.35 | 0.60 | 9 | -0.37 | 0.10 |
| ***Cortical ERPs*** |  |  |  |  |  |  |  |  |  |
| N1 Amplitude | 31 | 0.37 | -0.39 | 19 | 0.60 | -0.30 | 12 | -0.17 | -0.16 |
| LP Area | 31 | -0.56** | 0.37 | 19 | -0.51 | 0.35 | 12 | -0.47 | -0.04 |
| P300 Area | 27 | -0.48 | 0.13 | 18 | -0.45 | 0.19 | 9 | 0.00 | -0.73 |

\* $p < 0.05$ , \*\* $p < 0.01$

### Group Differences

Table 7 summarizes the measures obtained from the AM perception tasks for the age-matched TD and LiD samples that were used to examine all group differences. Since these groups were matched for age, it was controlled as a possible confound, and the statistics that were used to compare them did not employ age as a factor. The AM Threshold ANOVA yielded a significant main effect of modulation rate *F*(1,80) = 149.35, *p* < .001, η_p_^2^ = 0.65, in which thresholds at the 40 Hz modulation rate were lower (better) than at 4 Hz. There was also a significant main effect of group, in which thresholds for the TD group were lower than those for the LiD group, *F*(1,80) = 5.46, *p* = .022, η_p_^2^ = 0.06. These main effects were qualified by an interaction between modulation rate and group, *F*(1,80) = 6.56, *p* = .012, η_p_^2^ = 0.08. Posthoc analyses demonstrated that 40 Hz AM thresholds were better than 4 Hz thresholds within both groups (*p* < .001 in both cases). They also identified that the TD group had superior thresholds to the LiD group for 4 Hz (*p* = .012), but not 40 Hz AM (*p* = .489). Group differences were additionally apparent with respect to performance on the AM Change task, with the TD group exhibiting significantly better accuracy in the detection of alternations between 4 and 40 Hz AM, *t*(22) = 4.35, adj. *p* < .001, *d* = 1.78. There was no significant difference in RTs despite a large effect size for this contrast, *t*(22) = 2.06, adj. *p* = .103, *d* = 0.84. Performance for the two groups on the AM Threshold and AM Change tasks is illustrated in panel C of Figure 2.

**Table 7:** Descriptive statistics for group performance on all measures from the AM perception tasks.

|  | TD |  |  | LiD |  |  |
| --- | --- | --- | --- | --- | --- | --- |
|  | N | Mean (SD) | 95% CI | N | Mean (SD) | 95% CI |
| ***AM Threshold*** |  |  |  |  |  |  |
| 4 Hz (dB) | 41 | -12.02 (4.47) | -13.39, -10.66 | 41 | -9.57 (4.94) | -11.08, -8.06 |
| 40 Hz (dB) | 41 | -16.88 (3.19) | -17.86, -15.90 | 41 | -16.37 (4.78) | -17.84, -14.91 |
| ***AM Change Task*** |  |  |  |  |  |  |
| Accuracy (%) | 12 | 82.33 (14.62) | 74.06, 90.60 | 12 | 42.79 (27.86) | 27.03, 58.56 |
| Response Time | 12 | 778.98 (198.26) | 666.81, 891.15 | 12 | 969.37 (251.76) | 827.21, 1112.10 |
| ***ASSR SNR (dB re 1 μV)*** |  |  |  |  |  |  |
| Left Hemisphere | 14 | 5.48 (1.67) | 4.51, 6.44 | 14 | 5.38 (1.13) | 4.73, 6.03 |
| Right Hemisphere | 14 | 5.51 (1.58) | 4.59, 6.42 | 14 | 4.80 (1.36) | 4.02, 5.58 |
| 4 Hz | 14 | 4.36 (1.46) | 3.52, 5.20 | 14 | 4.07 (1.48) | 3.31, 4.92 |
| 40 Hz | 14 | 6.62 (2.13) | 5.39, 7.85 | 14 | 6.11 (1.40) | 5.30, 6.92 |
| ***ASSR Phase Coherence*** |  |  |  |  |  |  |
| Left Hemisphere | 14 | 0.64 (0.13) | 0.57, 0.72 | 14 | 0.66 (0.09) | 0.60, 0.71 |
| Right Hemisphere | 14 | 0.66 (0.11) | 0.59, 0.72 | 14 | 0.60 (0.12) | 0.53, 0.66 |
| 4 Hz | 14 | 0.57 (0.14) | 0.49, 0.65 | 14 | 0.54 (0.13) | 0.46, 0.62 |
| 40 Hz | 14 | 0.73 (0.14) | 0.66, 0.81 | 14 | 0.71 (0.12) | 0.64, 0.78 |
| ***Cortical ERPs*** |  |  |  |  |  |  |
| N1 Amplitude (μV) | 18 | -0.40 (1.53) | -1.11, 0.30 | 18 | 0.49 (1.23) | -0.08, 1.06 |
| LP Area (μVs) | 18 | 1.94 (1.69) | 1.16, 2.72 | 18 | 0.70 (0.49) | 0.47, 0.92 |
| P300 Area (μVs) | 9 | 1.85 (1.74) | 0.72, 2.99 | 9 | 1.31 (0.82) | 0.78, 1.84 |

The ANOVA for the SNR of the ASSR yielded a significant main effect of frequency, *F*(1,26) = 36.05, *p* < .001, η^2^_G_ = 0.28, in which the SNR tended to be higher for the 40 Hz than the 4 Hz ASSR. There were no main effects of either hemisphere, *F*(1,26) = 1.80, *p* = .192, η^2^_G_= 0.01, or group, *F*(1,26) = 0.63, *p* = .434, η^2^_G_ = 0.01. Similarly, there were no significant interactions between group and any other factor, namely frequency, *F*(1,26) = 0.09, *p* = .763, η^2^_G_ < 0.01, hemisphere, *F*(1,26) = 2.19, *p* = .151, η^2^_G_ = 0.01, or the three-way interaction between all of these factors, *F*(1,26) = 3.22, *p* = .084, η^2^_G_ = 0.01. There was also no significant interaction between frequency and hemisphere, *F*(1,26) = 0.30, *p* = .587, η^2^_G_ < 0.01.

The results of the ANOVA for phase coherence closely mirrored those for SNR. The only significant main effect was for frequency, *F*(1,26) = 31.90, *p* < .001, η^2^_G_= 0.26, with higher phase coherence for the 40 Hz than the 4 Hz ASSR. There was no significant main effect of group, *F*(1,26) = 0.428, *p* = .519, η^2^_G_ = 0.01, nor was there a significant interaction between group and frequency, *F*(1,26) = 0.01, *p* = .923, η^2^_G_ < 0.01, or a significant three-way interaction between group, frequency, and hemisphere, *F*(1,26) = 3.61, *p* = .069, η^2^_G_ = 0.01. There was also no significant interaction between frequency and hemisphere, *F*(1,26) = 0.133, *p* = .718, η^2^_G_ < 0.01. However, the interaction between hemisphere and group approached significance, *F*(1,26) = 4.20, *p* = .051, η^2^_G_ = 0.02. SNR and phase coherence metrics for the ASSR are illustrated as a function of frequency, group, and hemisphere in panel B of Figure 4.

Grand average waveforms for the ACC and P300 are shown in Figures 5 and 6, respectively. The N1 component of the ACC (upwards black arrow at E2 in Figure 5) was extremely small and did not cross baseline in the grand average. This tendency is also clearly reflected in the N1 amplitudes reported in Table 7. By contrast, the LP (downwards gray arrow at E23 in Figure 5) was visible for both groups and appeared markedly larger for the TD group. The P300 (indicated by a downwards gray arrow at E23 in Figure 6), which occurred in the same latency window as the LP, was similar for the two groups. There were no significant group differences in the amplitude of the N1, *t*(34) = 1.93, *p* = .062, *d* = 0.64, or the area of the P300, *t*(16) = 0.846, adj. *p* = .820, *d* = 0.40. The area under the LP curve, however, differed considerably between the two groups, *t*(19.83) = 3.01, adj. *p* = .014, *d* = 1.00.

**Figure 5.**
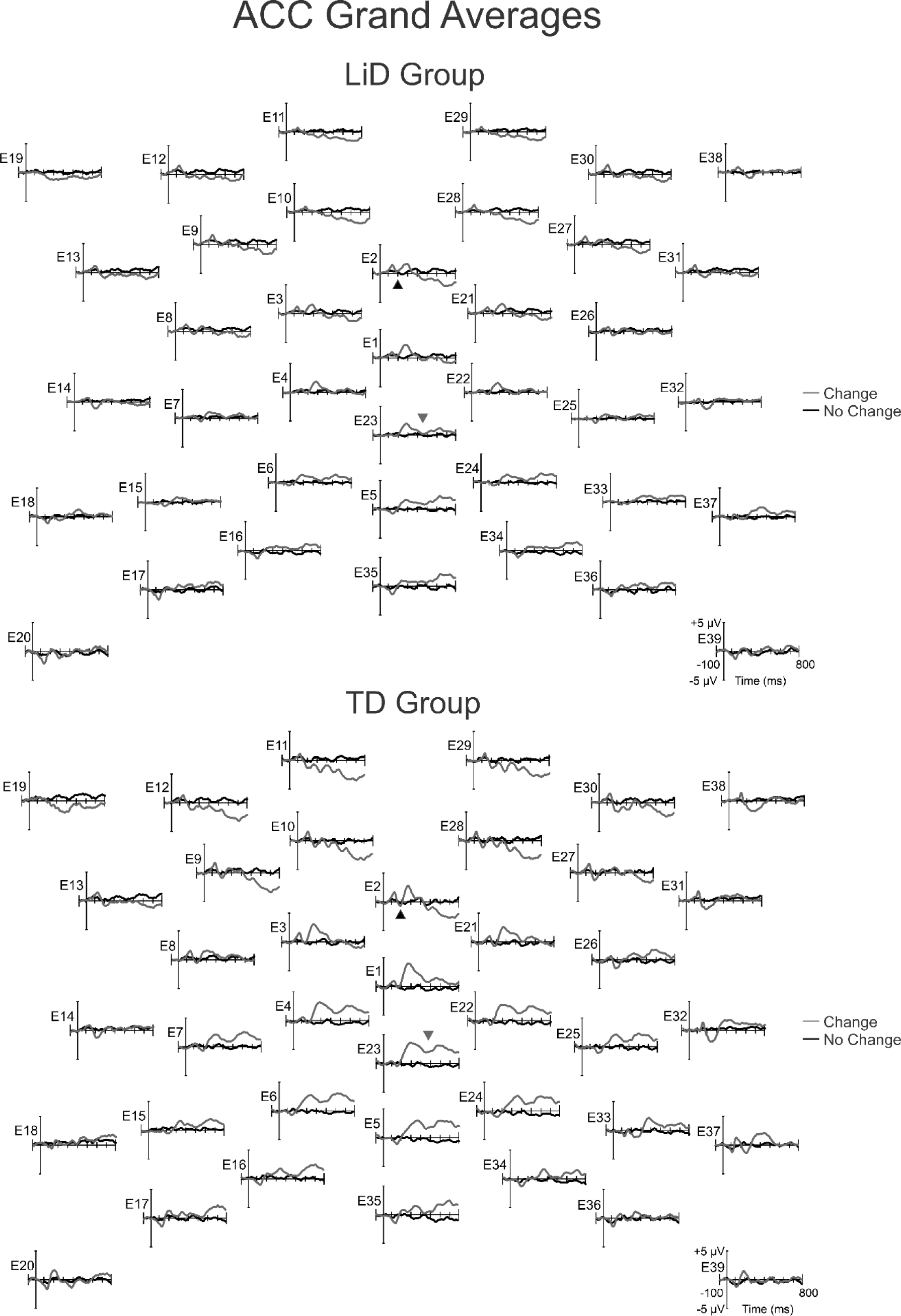
Grand average ACC waveforms as a function of group (LiD = top, TD = bottom) for the target (i.e., epochs containing an AM rate change; “Change” epochs), presented alongside the “No Change” (i.e., homogeneous AM) epochs. Black and gray arrows mark the latencies and locations of measurement for the N1 and LP, respectively. While N1 did not differ between the groups, there was a notable difference in the size of the LP. Note that the LP was quantified as an area under the curve, thus differences in the positioning of these arrows do not reflect differences in the period that was used for its measurement.

**Figure 6.**
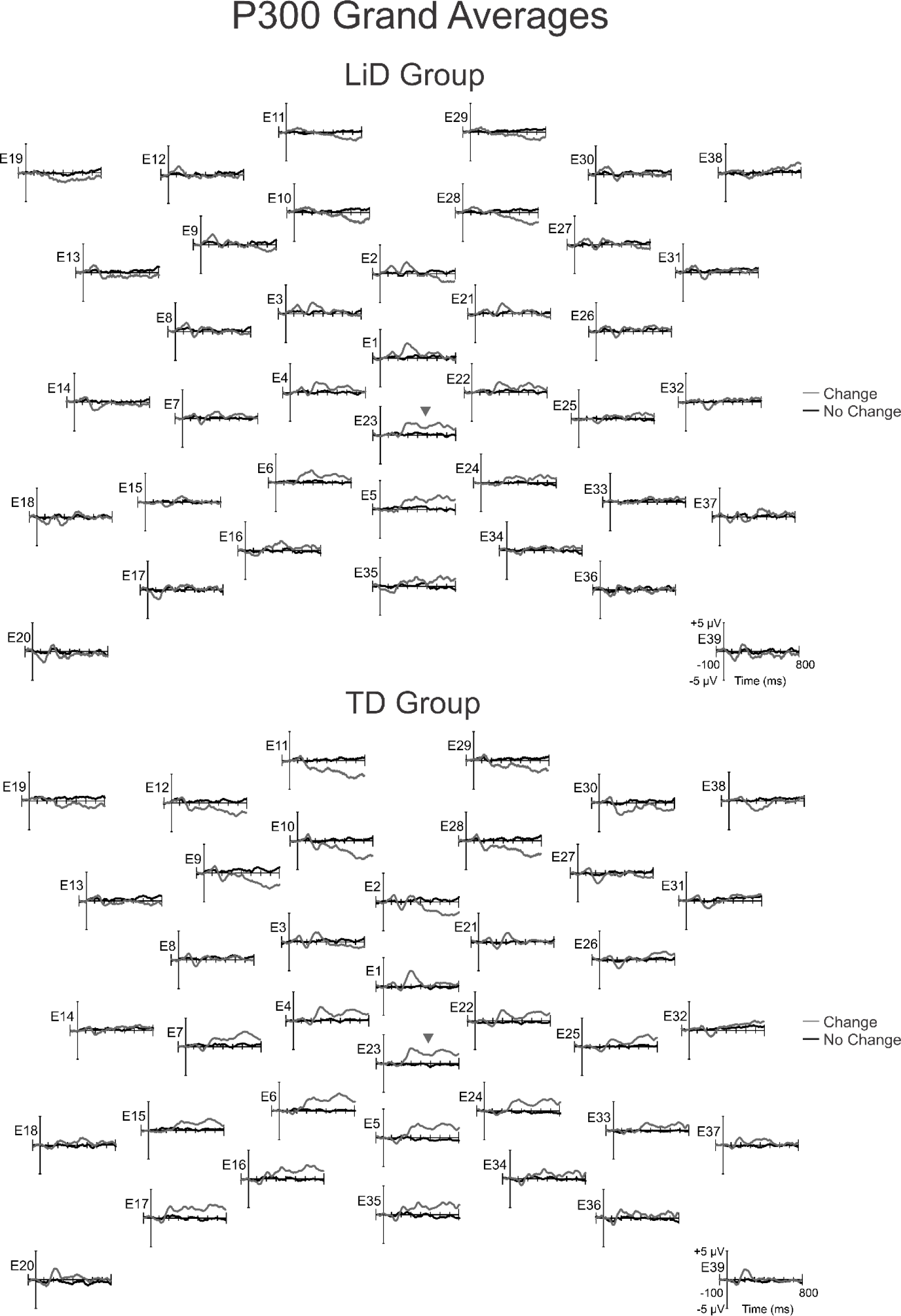
Grand average P300 waveforms as a function of group (LiD = top, TD = bottom) for correctly-detected target (i.e., “Change” epochs) vs. “No Change” (i.e., homogeneous AM) epochs. The location of the P300, which appeared as a complex, sustained positivity, is indicated by a gray arrow. Like the LP, the P300 was measured as an area under the curve. No group differences were observed for the P300.

## Discussion

Difficulty understanding SiN is a prominent feature of LiD and there is ample theoretical and experimental reason to suspect AM perception as a contributing factor to such deficits (Christiansen et al., 2013; Festen & Plomp, 1990; Gnansia et al., 2008; Horton et al., 2013). Nevertheless, few studies have investigated the relationship between AM perception and SiN task performance in children. For this reason, the first aim of this study was to provide such evidence, with age effects taken into account.

Maturation is an important consideration in the study of childhood auditory perceptual deficits, since there is considerable development of these skills into adolescence (Lopez-Poveda, 2014; Moore et al., 2008). In the present analysis, age exhibited significant correlations with performance in the Separated Talker condition of the CRM-based task, as well as 4 Hz AM thresholds, and the amplitude of the N1 component of the ACC, with older children demonstrating better thresholds and larger N1 responses. All of these effects are to be expected. Age-related increases in the amplitude of the N1 of the ACC have previously been observed (Martin et al., 2010) and both SiN performance and AM thresholds are known to improve over childhood (Cabrera et al., 2022; Hall & Grose, 1994; Talarico et al., 2007). The lack of such effects on the LiSN-S can be attributed to the fact that standard scores on this test are age-adjusted.

Overall, the correlations between measures of AM perception and SiN performance tended to be small. These relationships were distributed across a range of metrics. Significant correlations were obtained for both the 4 and 40 Hz modulation thresholds with the Separated Talker condition of the CRM-based task. AM thresholds at 4 Hz were also significantly correlated with performance in the Low Cue condition of the LiSN-S. In all cases, better AM task performance was associated with better SiN perception, providing some support for the notion that AM processing is an important ability for children to understand SiN. However, the within-group analyses suggested some possible disparities in these relationships as a function of LiD. Specifically, the relationship between 4 Hz AM thresholds and the Separated Talker condition of the CRM-based task may primarily have been driven by the TD group.

In 2020, Lotfi and colleagues also measured modulation thresholds (using spectrotemporal modulation) and SiN performance in children (separately for those with APD and their TD peers). In contrast to the present results, they found strong correlations across all temporal modulation rates and spectral modulation densities, in both groups. Any effort to compare the findings of these two studies must consider the differences in the SiN tasks that were used. Lotfi and colleagues employed very simple target speech (consonant vowel syllables and words) in the presence of maskers that provided limited informational masking (nonsense syllables and six-talker babble). By contrast, the present study employed single-talker sentences as both the target and masker. This is the type of task that children with LiD reportedly struggle with in school and at home – following meaningful verbal information from teachers and parents in the presence of other talkers. Some of the SiN measures used in the present study (i.e., the Talker and Spatial Advantage scores of the LiSN-S) were also subtractive scores, designed to isolate specific auditory skills (Cameron & Dillon, 2007). Given the fundamental differences in the tasks used for these studies, it is unsurprising that they yielded different results. Nevertheless, the present work supports and extends the argument that modulation sensitivity is related to SiN task performance in children.

One evoked response to the AM Change task, the LP, also correlated significantly with performance in the Separated Talker condition of the CRM-based task, with smaller LPs associated with poorer SiN performance. Similarly to the correlations involving 4 Hz AM thresholds, this effect may have primarily been driven by the TD group. The LP was measured in the same waveform as the ACC, which included all targets, regardless of whether they were accurately detected. As a result, smaller LPs could reflect genuine differences in the amplitude of the P300, a lower contribution of P300-containing epochs to the average due to inattention, or a combination of these factors.

The P300 is generally interpreted to reflect the updating of working memory following target detection (Polich, 2007), and in healthy individuals, modulations of its amplitude are related to the ease of perceptual discriminations (Polich, 1987), as well as cognitive abilities like intelligence (Amin et al., 2015) and working memory capacity (Daffner et al., 2011). While it is impossible to disentangle true changes in the size of the P300 from inattention in the LP, it is cautiously worth noting that the correlation between SiN performance and the area of the P300 (measured using only correctly-detected targets) did not reach significance. This might suggest that inattention is the source of smaller LPs for those who performed more poorly on the SiN task.

It may be worth noting that within-group analyses yielded medium effect sizes for some relationships that did not reach statistical significance in the across-group analysis. Specifically, the LiD group demonstrated a relationship between the age-adjusted amplitude of the N1 and Spatial Advantage scores on the LiSN-S, as well as between the SNR of the 40 Hz ASSR and performance in the Separated Talker condition of the CRM-based task. While these might be interpreted as evidence for sensory mechanisms for some SiN deficits, the small sample size involved here imposes a need for follow-up before drawing any such conclusions.

Another aim of the present study was to quantify the relationships between behavioral measures and evoked responses related to AM perception in children. This goal was important given that the inferential significance of the ACC N1 component with respect to AM sensitivity has only been established in adults (Han & Dimitrijevic, 2015, 2020). The observed correlations between behavioral measures and evoked responses tended to be modest. However, like the SiN correlations, these results again highlighted the importance of the LP, which correlated significantly with RT on the AM Change task, such that participants with larger LPs responded more quickly on the task. As with the SiN analysis, given the variety of factors that contribute to this response, it is difficult to make exact attributions regarding whether this reflects P300 modulation or inattention, though no such correlation was observed for the area of the P300.

Unlike the SiN analysis, the correlations between behavioral and evoked response measures of AM perception highlighted the importance of the 40 Hz ASSR. The SNR and phase coherence of the 40 Hz ASSR were significantly correlated with 4 Hz AM thresholds, and performance on the AM Change task (both accuracy and RT). Specifically, higher SNRs and phase coherence for the 40 Hz ASSR were associated with lower 4 Hz AM thresholds, higher accuracy on the AM Change task, and faster RTs. Although this pattern of results supports the notion that the ASSR is related to AM perception, the relative importance the 40 Hz ASSR is surprising.

Although the 40 Hz ASSR is an extremely robust response in adults, this is not always the case in pediatric populations. Infants between 3 weeks and 28 months of age show weak 40 Hz ASSRs (Stapells et al., 1988), and the response improves through adolescence (Cho et al., 2015; Rojas et al., 2006). This developmental pattern invites the possibility that, in children, the 40 Hz ASSR reflects both synchronization to the stimulus (i.e., sensory processing of AM) and maturation. Thus, it may be that participants with more mature 40 Hz ASSRs tended to perform better on behavioral metrics of AM perception. This interpretation is complicated by the fact that the correlation between age and these measures of the 40 Hz ASSR did not reach statistical significance in the across-group analysis. This failure may reflect the low power afforded by our sample. However, it may be worth noting that the effect of age on the 40 Hz ASSR appeared to be somewhat more robust for children in the LiD group [SNR *r_s_*(14) = 0.36, phase coherence *r_s_*(14) = 0.31] than those in the TD group [SNR *r_s_*(26) = 0.24, phase coherence r_s_(26) = 0.15]. Thus, it remains possible that this developmental process was more evident for children with LiD than their TD peers. This may also explain the observed relationship between the SNR of the 40 Hz ASSR and performance in the Separated Talker condition of the CRM-based task for children in the LiD group, since this SiN measure also varied with age. Regardless of the underlying reason for the relative importance of the 40 Hz ASSR in the present results, it is clear that, in children, there are no straightforward relationships between ASSRs to AM at 100% modulation depths and AM detection thresholds. They also do not support using the N1 of the ACC as an objective index of AM detection thresholds in children, though such a role was suggested based on measurements in adults (Han & Dimitrijevic, 2015, 2020).

The within-group correlations also yielded a medium-large effect size for a relationship between the area of the P300 and accuracy on the AM Change task, only for children with LiD. The observed direction of the effect, in which smaller P300s were associated with higher accuracy, was inconsistent with the typical finding that the P300 is larger for easier discriminations (Polich, 1987). In addition to task difficulty, the amplitude of the P300 is modulated by the target-to-target interval, with less frequent targets evoking larger P300s (Croft et al., 2003; Ladish & Polich, 1989; Polich, 1987). Since accuracies tended to be poor for children in the LiD group, many of the target events went undetected, perhaps rendering them subjectively less frequent. However, given the very small sample of children with LiD who could be used for the P300 analysis (N = 9), these results must be interpreted with extreme caution.

The ultimate goal of this analysis was to explore the basis of AM perception deficits in LiD using behavioral performance and evoked responses. Unlike the correlational analyses, these comparisons were performed using age-matched groups. The LiD group demonstrated poorer performance on AM perception tasks, including higher 4 Hz AM detection thresholds and lower accuracy on the AM Change task. The observation of deficits with 4 Hz but not 40 Hz AM may be surprising, since some studies have demonstrated lower sensitivity to higher rate AMs in both children and adults (Hall & Grose, 1994). However, the present data showed the opposite effect, with lower thresholds for both groups at 40 Hz than 4 Hz. Since the durations of the stimuli (1 second) were held constant across these two rates, the 4 Hz AM stimuli offered participants fewer cycles across which to detect modulation. Some lines of research suggest that children perform poorly compared to adults with stimuli that provide few cycles of AM due to inefficiencies in echoic memory (Cabrera et al., 2022). Modulations at this rate also have greater theoretical relevance for speech perception, since speech envelopes tend to modulate between 2 and 10 Hz (Ding et al., 2017). Thus, AM detection thresholds at 4 Hz may have special significance for LiD. Within-group effect sizes suggested that the relationship between 4 Hz AM thresholds and SiN performance may be stronger in TD children than those with LiD. This finding might implicate a mechanism for SiN deficits in LiD that involves impaired echoic memory.

Evoked responses were also used to provide insight into the auditory and cognitive processes underlying AM perception deficits with LiD. Specifically, the ASSR directly reflects the envelope phase-locked activity of sensory neurons in the brainstem and auditory cortex (Herdman et al., 2002). Some research suggests the existence of additional cortical non-auditory sources, potentially supporting multisensory integration (Farahani et al., 2021). No significant differences were observed with respect to the ASSR at either 4 or 40 Hz between children with and without LiD. Indeed, the only significant effect with respect to either SNR or phase coherence was a main effect of frequency, in which 40 Hz ASSRs exhibited higher SNRs and phase coherence than 4 Hz responses. While the relative SNR of the ASSR has not been systematically explored across AM rates in children, one study using transient tonal stimuli in children and adults demonstrated challenges with ASSR measurement at lower rates of stimulation due to a shift of power towards the harmonics of the stimulation rate, and increased noise (Tlumak et al., 2012). The present results are consistent with that finding.

On the basis of theories that propose hemispheric specializations for modulation processing (Poeppel, 2003), one line of research has explored the link between lateralization of the ASSR and dyslexia in children (Vanvooren et al., 2014). Though they found no differences in lateralization, dyslexia shares many symptoms with APD and auditory processing deficits may contribute to its pathophysiology (Dawes & Bishop, 2009, 2010). Thus, the possibility of altered lateralization was considered in the present study by including hemisphere as a factor in both ANOVAs for the ASSR. There was little evidence for any differences in lateralization between children who have LiD and their TD peers, except a near-significant interaction between hemisphere and group for phase coherence of the ASSR, with a very small effect size (η^2^_G_= 0.02). As such, like dyslexia, abnormal hemispheric specializations for AM processing do not appear to be a contributing factor in LiD.

The AM Change task also permitted the measurement of the ACC N1 component, the P300, and a LP in the same latency range as the P300. As previously reviewed, the ACC does not require attention to be evoked and corresponds well with behaviorally-measured discrimination thresholds (He et al., 2012; J.-R. Kim, 2015). However, despite considerable group differences in 4 Hz AM thresholds and accuracy on the AM Change task, there were no group differences in N1 amplitude. This may be attributable to the fact that this response was small and inconsistent in the present sample.

While the ASSR and ACC reflect sensory-perceptual operations, the P300 and LP index cognitive stages of processing. Reductions in the amplitude of the P300 have been interpreted as evidence of altered cognitive function in a wide range of clinical conditions (e.g., concussion, Petley et al., 2018; schizophrenia, Bramon, 2004; and dementia Hedges et al., 2016; among many others). However, this response did not yield significant differences between the LiD and TD groups. By contrast, there were large group differences in the size of the LP. This effect is readily evident in Figure 5 and Table 7. A large, multi-peaked, slow LP dominated the response to target stimuli at parietal sites from 200 ms onwards for TD participants, yet only a small, transient LP with very little sustained activation was seen for children with LiD. Statistically, the group difference in the area under the LP curve had a large effect size (*d* = 1.00). As with the previously-observed correlations between the area under the LP curve and measures of SiN and AM perception, it is difficult to make a precise statement regarding the implications of this effect. The lack of a group difference in the P300, however, might suggest that it reflects inattention. Regardless of its specific meaning, the effect is undoubtedly cognitive in nature, and reflects the activity of brain regions that lie outside the CANS.

All tests of central auditory processing are influenced by cognition, and obtaining measures that are relatively free from such factors requires targeted strategies in test design (Moore & Dillon, 2018). It should not be surprising, therefore, that measures of AM perception might, at least in part, reflect cognitive factors. Perception and cognition also interact considerably, to the extent that impaired perceptual object formation could itself result in poorer selective attention (Shinn-Cunningham, 2008). In this way, any study that aims to identify the stage of processing at which perceptual deficits occur relies on a somewhat artificial distinction. Nevertheless, evoked responses offer the opportunity for more specificity in such inferences than behavior alone. The results of the present study confirm that AM perception is important for SiN performance in children, and that a pediatric population that is susceptible to SiN perception deficits – children with LiD – is impaired with respect to this ability. Correlational analyses raised the possibility that cortical development, as reflected by the 40 Hz ASSR, and echoic memory may contribute to AM and SiN perception in this group. The evoked responses that characterized LiD, however, consisted only of a reduced cognitive response during AM change detection, which may suggest a primarily cognitive origin for these deficits.

## Data Availability

All data produced in the present study are available upon reasonable request to the authors after publication

## Acknowledgements

This research was supported by grant R01 DC014078 from the National Institute on Deafness and Other Communication Disorders, and by the Cincinnati Children’s Research Foundation. DRM is also supported by the NIHR Manchester Biomedical Research Centre. The authors gratefully acknowledge the assistance of A. Dimitrijevic, N. Sloat, A. Perdew, and N. Clevenger. We also thank the families who supported this project through participation as well as the Summer Undergraduate Research Foundation (SURF) scholars who assisted with the project.

## Data Availability

Data will be made available on request to the authors.

## Supplementary Materials

**Supplementary Table 1.** A summary of EEG denoising and ERP data quality metrics.

