## Supplemental Table 1 for "Amplitude modulation perception and cortical evoked potentials in children with listening difficulties and their typically-developing peers"

Supplementary Table 1: EEG denoising and ERP data quality metrics. Number of ICA components rejected and number of trials accepted for averaging are reported as mean *(SD)*. Measurement windows for ERP amplitudes and areas under the curve are reported as window limits, in milliseconds.

|  | **TD** | **LiD** |
| --- | --- | --- |
| ***ICA Components Rejected*** | 3.4 *(0.9)* | 3.5 *(0.9)* |
| ***Trials Accepted for Averaging*** |  |  |
| *ACC (No Change)* | 217.9 *(35.6)* | 216.6 *(28.5)* |
| *ACC (Change)* | 210.4 *(35.0)* | 205.6 *(36.4)* |
| *P300 (No Change)* | 220.9 *(36.1)* | 210.1 *(33.3)* |
| *P300 (Correctly Detected Changes)* | 160.1 *(49.3)* | 96.9 *(39.7)* |
| ***ERP Measurement Window*** |  |  |
| *N1* | 143 – 183 | 139.5 – 179.5 |
| *LP* | 156.0 – end | 180.5 – end |
| *P300* | 184.5 – end | 195.0 – end |
